# Diagnostic Codes in AI prediction models and Label Leakage of Same-admission Clinical Outcomes

**DOI:** 10.1101/2025.08.09.25333360

**Authors:** Bashar Ramadan, Ming-Chieh Liu, Michael C. Burkhart, William F. Parker, Brett K. Beaulieu-Jones

## Abstract

**Importance:** Artificial intelligence (AI) and statistical models designed to predict same-admission outcomes for hospitalized patients, such inpatient mortality, often rely on International Classification of Disease (ICD) diagnostic codes, even when these codes are not finalized until after hospital discharge.

**Objective:** Investigate the extent to which the inclusion of ICD codes as features in predictive models inflates performance metrics via “label leakage” (e.g. including the ICD code for cardiac arrest into an inpatient mortality prediction model) and assess the prevalence and implications of this practice in existing literature.

**Design:** Observational study of the MIMIC-IV deidentified inpatient electronic health record database and literature review.

**Setting:** Beth Israel Deaconess Medical Center.

**Participants:** Patients admitted to the hospital with either emergency room or ICU between 2008 and 2019

**Main outcome and measures:** Using a standard training-validation-test split procedure, we developed multiple AI multivariable prediction models for inpatient mortality (logistic regression, random forest, and XGBoost) using only patient age, sex, and ICD codes as features. We evaluated these models in the test set using area under the receiver operating curves (AUROC) and examined variable importance. Next, we determined the percentage of published multivariable prediction models using MIMIC that used ICD codes as features with a systematic literature review.

**Results:** The study cohort consisted of 180,640 patients (mean age 58.7 ranged from 18-103, 53.0% were female) and 8,573 (4.7%) died during the inpatient admission. The multivariable prediction models using ICD codes predicted in-hospital mortality with high performance in the test dataset (AUROCs: 0.97-0.98) across logistic regression, random forest, and XGBoost. The most important ICD codes were ‘brain death,’ ‘cardiac arrest’, ‘Encounter for palliative care’, and ‘Do Not resuscitate status’. The literature review found that 40.2% of studies using MIMIC to predict same-admission outcomes included ICD codes as features even though both MIMIC publications and documentation clearly state the ICD codes are derived after discharge.

**Conclusions and relevance:** Using ICD codes as features in same-admission prediction models is a severe methodological flaw that inflates performance metrics and renders the model incapable of making clinically useful predictions in real-time. Our literature review demonstrates that the practice is unfortunately common. Addressing this challenge is essential for advancing trustworthy AI in healthcare.

**Key Points:** *Question:* Do International Classification of Disease (ICD) diagnostic codes, which are only finalized after hospital discharge, artificially inflate the performance of AI healthcare prediction models?

*Findings:* In a systematic literature review, 40.2% of published models trained to predict same-admission outcomes on the benchmark MIMIC dataset use ICD codes as features, despite both MIMIC papers clearly stating these codes are only available after discharge. Prediction models for inpatient mortality trained on ICD codes alone in the MIMIC-IV dataset can predict in-hospital mortality with high accuracy (AUROCs: 0.97-0.98). The most important codes are not available in time for any clinically useful mortality prediction (e.g. “brain death” and “Encounter for palliative care”).

*Meaning:* ICD codes are frequently used in inpatient AI prediction models for outcomes during the same admission rendering their output clinically useless. To ensure AI models are both reliable and clinically deployable, greater diligence is needed in identifying and preventing label leakage.

## Introduction

AI/ML models have demonstrated impressive performance in predicting critical same-admission outcomes, such as in-hospital mortality.^1–3^Some models use International Classification of Disease (ICD) diagnostic billing codes as input features. Since ICD codes are entered in the record *after* a clinical event, can be revised over the course of an admission, and are finalized only after discharge, their inclusion introduces data leakage, where information unavailable in real-world settings is improperly used during model training and evaluation.

For example, imagine a patient admitted with ‘unspecified abdominal pain’. After further evaluation, the patient is diagnosed with appendicitis, develops septic shock, and several days later suffers cardiac arrest before passing away. Early in the patient’s admission, only ‘unspecified abdominal pain’ would be available. However, if a model incorporates all ICD codes subsequently assigned after the end of a hospital stay, it unfairly leverages hindsight information to predict mortality, achieving deceptively high accuracy.

To illustrate the impact of this problem, we performed two analyses. First, we use ICD codes in models predicting inpatient mortality, one of the most common same-admission prediction tasks. Second, we performed a systematic review of research papers that build AI models to predict inpatient outcomes and identified the percentage of those that included ICD codes from that admission as input features. The potential for label leakage has been previously described^4^, and there are a wide array of published examples of “shortcut learning” in machine learning for healthcare^5–9^. This work extends further to clearly illustrate the impact of label leakage on a common same admission prediction task and to quantify the level at which label leakage exists in the literature.

## Methods

### Data source and study population

We used the Medical Information Mart for Intensive Care IV database version 2.2 (MIMIC-IV v2.2) a publicly available deidentified electronic healthcare record database of patients admitted to an ICU or emergency department at Beth Israel Deaconess Medical Center between 2008 and 2019. All admissions with ICD codes were included in our study, with less than 1% excluded. We partitioned the dataset by the date of admission into train (70%), validation (10%), test (20%) sets per TRIPOD-AI+ guidelines,^10^ excluding patients from the validation and test sets who also had admissions in the training set. During pre-processing, we converted ICD-10 codes to ICD-9 and removed ICD codes that had low variance (<0.0001) or high covariance (>0.8) with other ICD codes. The features “age” and “sex” remained in the models.

### ICD code prediction model development and evaluation

We trained classification models (logistic regression^11^, random forest^11^, and XGBoost^12^) using only patient age, sex, and ICD-9 codes as features, tuning hyperparameters in validation set. We chose these models as they are some of the commonly used classifiers, achieve strong performance with tabular data, and offer approaches to interpret models. Other predictive features such as vital signs, lab values, and medications were intentionally excluded to examine only the potential for ICD code-driven label leakage. The trained models were then evaluated on the held-out test set, with performance assessed using AUROC and balanced accuracy. We calculated odds ratios and p-values for ICD codes in the logistic regression model and applied the Benjamini–Hochberg procedure to control for false discovery rate with a threshold of p < 0.05. For the random forest and XGBoost models, we assessed feature importance with each library’s respective default criterion, namely Gini importance and gain, to identify key ICD codes for the prediction task.

Full source code is available on Github (https://github.com/bbj-lab/data-leakage).

### Systematic literature review

We employed Google Scholar in November 2024 with two search queries: (1) “prediction model machine learning “mimic-IV” OR “mimic IV” OR “mimic 4” OR “mimic-4”“ and (2) “prediction model machine learning “mimic-III” OR “mimic III” OR “mimic 3” OR “mimic-3”.” We sorted results by citations per year to avoid bias against recently published studies and screened sequentially until we identified 100 predictive modeling studies (50 each from MIMIC-III and MIMIC-IV). We categorized these papers by whether they predicted clinical events during the same admission and whether ICD codes from that admission were included as input features.

## Results

### ICD code-based prediction models

The study cohort included 422,534 hospital admissions from 180,640 unique patients. The average age at admission was 58.69 years with standard deviation 19.23 years. Among the patients, 84,965 were male. In-hospital mortality occurred in 8,417 admissions. In the held-out test set, all three models achieved high predictive performance, with AUROCs of 0.98 (logistic regression), 0.97 (random forest), and 0.97 (XGBoost) (**Figure 1A, eTable 1**). These results are even better than published models trained on the same data that also included many additional predictive features from the rest of the electronic medical record.^1,2^

**Figure 1.**
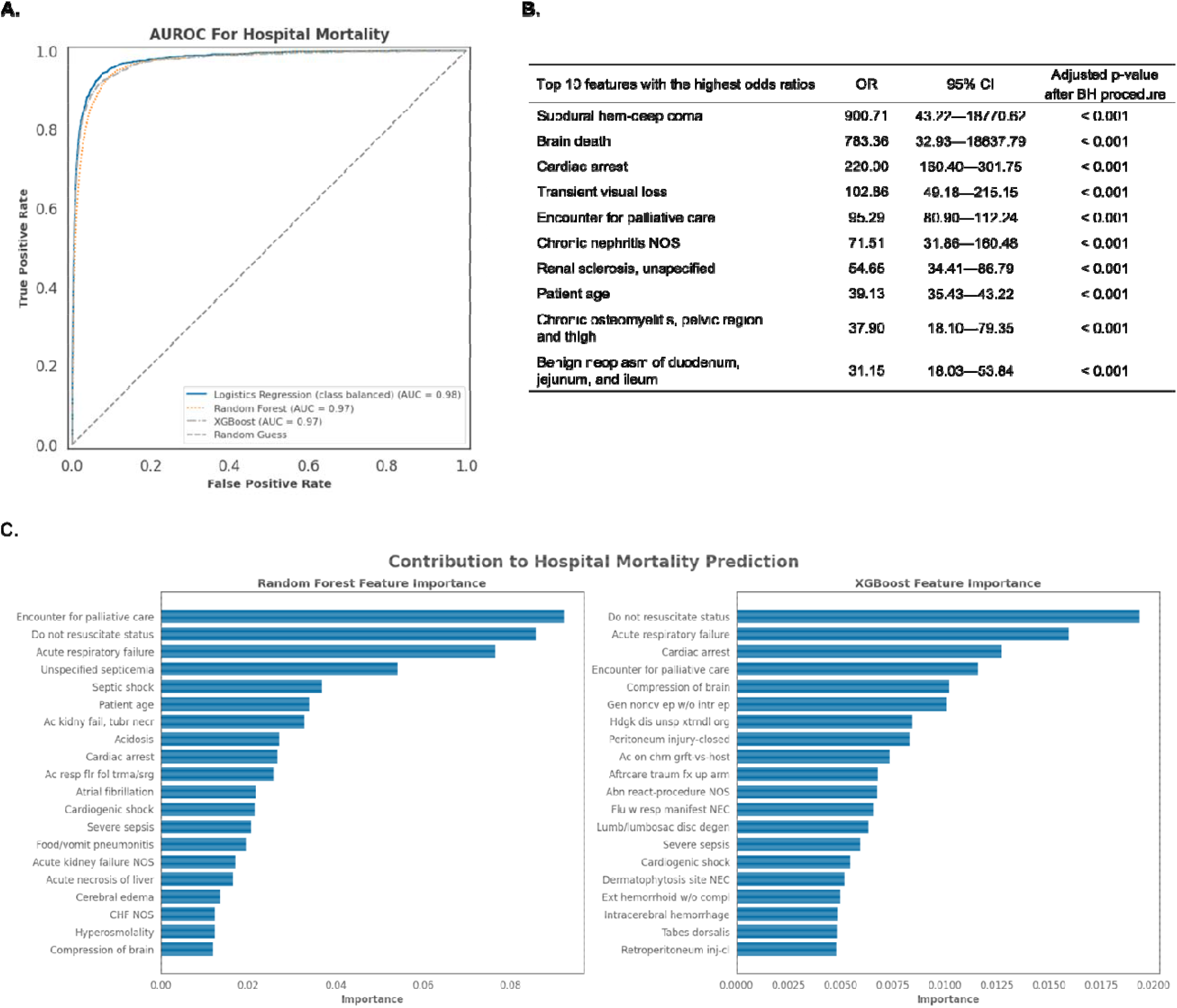
A.) AUROC of logistics regression models, random forest model, and XGBoost model for mortality prediction task in the hold-out test set, **B.)** Selected features correlated to mortality from logistics regression analysis, **C.)** Feature importance for random forest model and XGBoost model predicting mortality.

**Figure 1B** highlights significant diagnostic codes used by the logistic regression model with the adjusted p-values <0.05 after the Benjamini–Hochberg procedure. Acute diagnoses typically arising during hospitalization dominate the list. These include diagnoses such as ‘subdural hematoma – deep coma’, ‘brain death,’ and ‘cardiac arrest’ all of which carry an obvious high risk of mortality. Feature importance analyses from the random forest and XGBoost models (**Figure 1C**) found ICD codes ‘Do not resuscitate’ status, ‘acute respiratory failure’ and ‘encounter for palliative care’ to be powerful predictors of mortality.

In addition to ICD codes that obviously represent label leakage (e.g. ‘brain death’), the diagnosis ‘external hemorrhoids without complications’ was important to random forest predictions which stands out as it is not an acute diagnosis. This anomaly may reflect the model’s ability to detect a clinician’s focus on documenting less severe conditions, signaling relative patient stability and, therefore, low mortality risk.

### Literature review

Figure 2. outlines our paper-screening process. We reviewed 100 papers building a predictive model from an initial set of the 139 papers citing MIMIC sorted in descending order by the average number of citations per year (Full list in **eTable 2**). Of these, 92 built predictive models targeting outcomes within the same admission, and among them, 40.2% (37/92) used ICD diagnostic codes as input features.

## Discussion

Our findings elucidate a specific but pervasive problem within the machine learning healthcare literature: the presence of data leakage in same-admission prediction models caused by the inclusion of diagnostic codes as input features. These codes, finalized only after discharge, provide models with hindsight information that would not be available at the time of prediction. This practice causes two distinct and serious problems. First, codes that clinicians document in the electronic health record after a clinical encounter cannot be used to guide real-time clinical decision making during that encounter. Second, a subset of these codes (e.g. brain death for inpatient mortality) document highly correlated events with the outcome being predicted. This issue underscores a broader concern: machine learning models trained with retrospective data risk misrepresenting their value in real-world clinical care. If these models fail to account for the realities of real-time clinical workflows, their success in research will not translate into meaningful improvements in patient outcomes.

**Figure 2.**
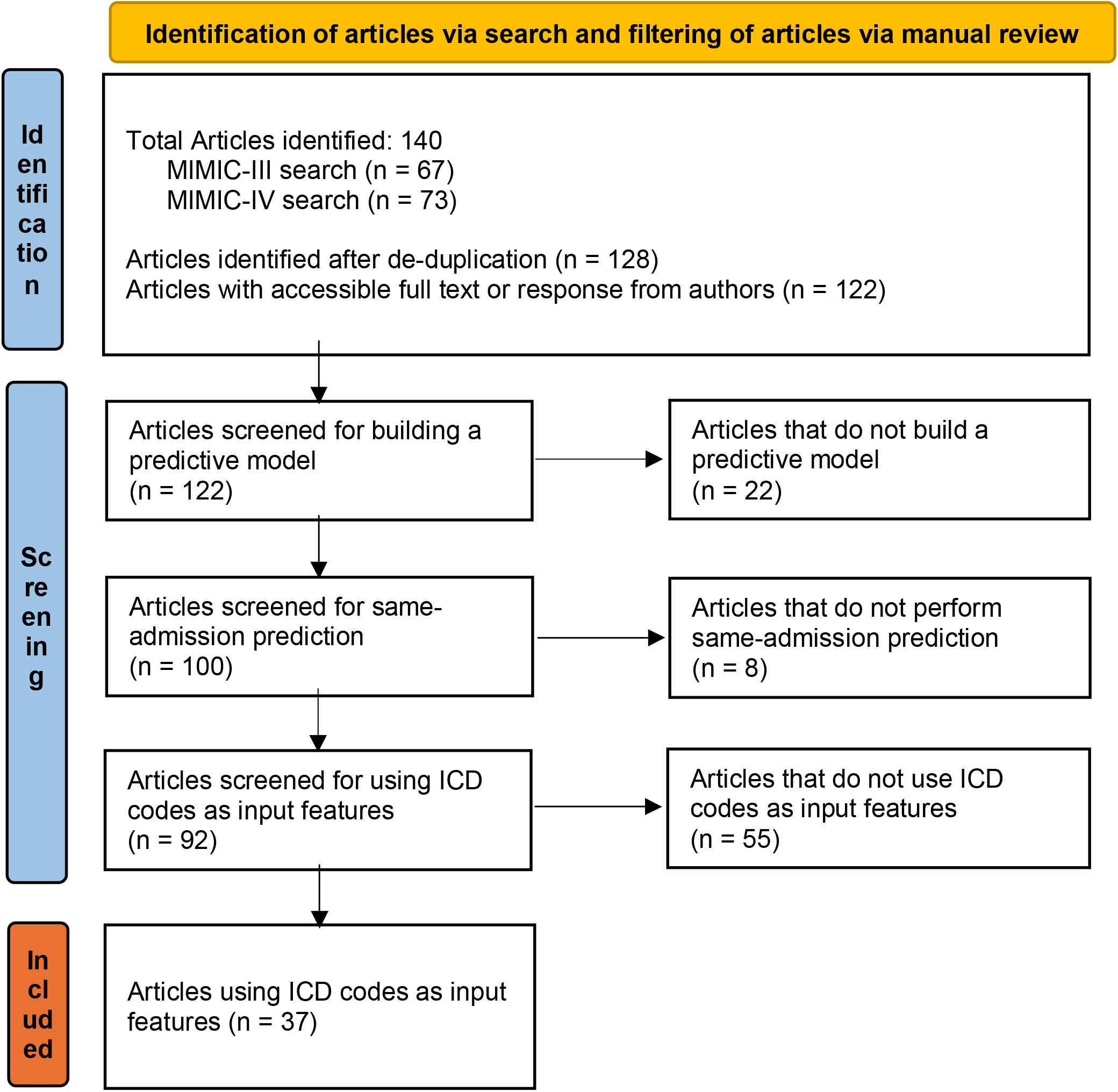
Search and filtering process of articles citing MIMIC-III or MIMIC-IV, developing a prediction model, performing same admission predictions, and using ICD codes as input features.

Both MIMIC-III and MIMIC-IV carry explicit warnings against using an admission’s ICD codes to predict outcomes from that same admission. In MIMIC-III, ICD-9 codes arise “from patient discharges,”^13^ while MIMIC-IV clarifies that diagnoses are determined “by trained professionals after reviewing signed patient notes.”^14^ These datasets do not provide an “audit” log of changes or updates to ICD codes, but instead provide only the final set of ICD diagnoses. Given the prevalence of ICD code use in MIMIC based studies despite this direct guidance, this leads us to suspect that publications on private institutional data, especially those that do not share source code, may be even more likely to be compromised by label leakage.

Researchers aim to harness available knowledge to the greatest extent possible when training models, and there’s a reasonable expectation that some diagnoses are known to clinicians shortly after admission, e.g. broken limbs and burns. Some information could potentially be gleaned from patient notes or physician problem lists that may be available during a patient’s stay. Oftentimes codes are carried over from previous visits, e.g. chronic conditions or comorbidities such as diabetes and hypertension, and these can safely be assumed known.

However, diagnoses in the form of ICD codes for a given admission in MIMIC are explicitly derived after discharge. In other datasets it may be possible to use ICD codes without label leakage if these codes are timestamped and derived from problem lists. However, there are still substantial limitations given the fact that these codes are used for billing purposes and represent clinical thinking as opposed to patient state^3^.

Both analyses in our study are limited because they only the benchmark MIMIC dataset. However, thousands of papers have relied on data from the MIMIC database^13,14^ for clinical prediction tasks, and a significant portion incorporate ICD codes to predict same-admission outcomes. It is very unlikely that this problem is isolated to MIMIC database work but reflects a broader challenge in healthcare machine learning research^4^. The fact that label leakage occurs this often in a well-defined dataset which explicitly describes the nature of ICD codes should raise pause for research using less transparent datasets and methods.

The solution to these problems, like solutions to other instances of label leakage, data leakage, and shortcut learning are to diligently examine the input features and to analyze what models are relying on for their output decisions. The utility of ICD codes geared at billing for deployable predictive models is debatable but at a minimum, researchers need to be careful to ensure the codes are available prior to the time a prediction needs to be made. This may require only using codes from prior admissions, which still requires ensuring they are not edited during any adjudication processes with payors or deriving these diagnoses from a timestamped problem list. MIMIC, however, does not include either timestamps or codes from the problem list. The frequency of this error indicates a need for researchers to more closely read the documentation of third-party datasets. While it is not possible to estimate how frequently this occurs on private, institutional datasets, we believe the frequency also indicates a need for greater engagement of predictive model developers with experts covering the full data generation (clinicians) and preparation (e.g., data warehousing-IT) process.

## Conclusion

Using ICD codes as features in same-admission prediction models is a widespread practice that produces AI models that appear effective in research settings but could never be deployed in real-world clinical environments. Addressing this challenge is essential for advancing trustworthy AI in healthcare.

## Data Availability

Data used in this study are available in the supplemental materials (literature review) and via Physionet (https://physionet.org/).

