## Supplemental Information for "Diagnostic Codes in AI prediction models and Label Leakage of Same-admission Clinical Outcomes"

**Supplementary information**

**eTable 1. Full list of features’ odds ratios for logistics regression model.** Supplementary file supplemental_data*.xlsx* gives the full list of significance of features in the logistics regression model.

**eTable 2. Papers citing MIMIC which build a predictive model.**

| Title | Pub status | Mimic 3 v 4 | Same admission prediction? | ICD codes used as features? |
| --- | --- | --- | --- | --- |
| Predicting 30-days mortality for MIMIC-III patients with sepsis-3: a machine learning approach using XGboost^1^ | Journal | III | FALSE | FALSE |
| Prediction model of in-hospital mortality in intensive care unit patients with heart failure: machine learning-based, retrospective analysis of the MIMIC-III database^2^ | Journal | III | TRUE | TRUE |
| Predicting sepsis with a recurrent neural network using the MIMIC III database^3^ | Journal | III | TRUE | FALSE |
| Machine-learning models for prediction of sepsis patients mortality^4^ | Journal | IV | TRUE | FALSE |
| Machine learning prediction models for prognosis of critically ill patients after open-heart surgery^5^ | Journal | III | FALSE | TRUE |
| A machine learning-based prediction model for in-hospital mortality among critically ill patients with hip fracture: An internal and external validated study^6^ | Journal | III | TRUE | TRUE |
| Machine learning prediction models for mechanically ventilated patients: analyses of the MIMIC-III database^7^ | Journal | III | TRUE | TRUE |
| Towards a decision support tool for intensive care discharge: machine learning algorithm development using electronic healthcare data from MIMIC-III and Bristol, UK^8^ | Journal | III | TRUE | FALSE |
| Development and validation of a machine-learning model for prediction of extubation failure in intensive care units^9^ | Journal | IV | TRUE | FALSE |
| Benchmarking emergency department prediction models with machine learning and public electronic health records^10^ | Journal | IV | TRUE | FALSE |
| Application of interpretable machine learning for early prediction of prognosis in acute kidney injury^11^ | Journal | IV | TRUE | TRUE |
| Developing an explainable machine learning model to predict the mechanical ventilation duration of patients with ARDS in intensive care units^12^ | Journal | IV | TRUE | FALSE |
| Developing machine learning models for prediction of mortality in the medical intensive care unit^13^ | Journal | III | TRUE | FALSE |
| A novel machine learning model to predict respiratory failure and invasive mechanical ventilation in critically ill patients suffering from COVID-19^14^ | Journal | III | TRUE | FALSE |
| Predicting hospital length of stay using neural networks on mimic iii data^15^ | Journal | III | TRUE | TRUE |
| A machine learning-based prediction model for acute kidney injury in patients with congestive heart failure^16^ | Journal | III | TRUE | FALSE |
| Explainable machine-learning model for prediction of in-hospital mortality in septic patients requiring intensive care unit readmission^17^ | Journal | IV | TRUE | FALSE |
| Atrial fibrillation detection during sepsis: study on MIMIC III ICU data^18^ | Journal | III | TRUE | FALSE |
| A machine-learning approach for dynamic prediction of sepsis-induced coagulopathy in critically ill patients with sepsis^19^ | Journal | IV | TRUE | TRUE |
| Prediction of intensive care unit length of stay in the MIMIC-IV dataset^20^ | Journal | IV | TRUE | TRUE |
| Early prediction of ventilator-associated pneumonia in critical care patients: a machine learning model^21^ | Journal | III | TRUE | FALSE |
| Predicting duration of mechanical ventilation in acute respiratory distress syndrome using supervised machine learning^22^ | Journal | III | TRUE | FALSE |
| Prediction of length-of-stay at intensive care unit (icu) using machine learning based on mimic-iii database^23^ | Journal | III | TRUE | TRUE |
| Mapping patient trajectories using longitudinal extraction and deep learning in the MIMIC-III critical care database^24^ | Journal | III | TRUE | FALSE |
| Machine learning prediction models and nomogram to predict the risk of in-hospital death for severe DKA: A clinical study based on MIMIC-IV, eICU^25^ | Journal | IV | TRUE | TRUE |
| Development of a machine learning-based prediction model for sepsis-associated delirium in the intensive care unit^26^ | Journal | IV | TRUE | TRUE |
| Novel pneumonia score based on a machine learning model for predicting mortality in pneumonia patients on admission to the intensive care unit^27^ | Journal | IV | TRUE | TRUE |
| Predictive modeling in urgent care: a comparative study of machine learning approaches^28^ | Journal | III | TRUE | TRUE |
| Development of a nomogram to predict 28-day mortality of patients with sepsis-induced coagulopathy: an analysis of the MIMIC-III database^29^ | Journal | III | TRUE | FALSE |
| Mortality prediction for patients with acute respiratory distress syndrome based on machine learning: a population-based study^30^ | Journal | III | TRUE | FALSE |
| Machine learning models for early prediction of sepsis on large healthcare datasets^31^ | Journal | III | TRUE | FALSE |
| Outcome Prediction in Critically-Ill Patients with Venous Thromboembolism and/or Cancer Using Machine Learning Algorithms: External Validation and Comparison with Scoring Systems^32^ | Journal | III | TRUE | TRUE |
| Statistical analysis and machine learning prediction of disease outcomes for COVID-19 and pneumonia patients^33^ | Journal | III | TRUE | FALSE |
| Prediction model of in-hospital mortality in intensive care unit patients with cardiac arrest: a retrospective analysis of MIMIC-IV database based on machine^34^ | Journal | IV | TRUE | TRUE |
| Development and assessment of scoring model for ICU stay and mortality prediction after emergency admissions in ischemic heart disease: a retrospective study of MIMIC-IV databases^35^ | Journal | IV | TRUE | TRUE |
| Tendency of dynamic vasoactive and inotropic medications data as a robust predictor of mortality in patients with septic shock: An analysis of the MIMIC-IV database^36^ | Journal | IV | TRUE | FALSE |
| Benchmarking PySyft federated learning framework on MIMIC-III dataset^37^ | Journal | III | TRUE | FALSE |
| Development and validation of a deep learning model to predict the survival of patients in ICU^38^ | Journal | III | TRUE | FALSE |
| Establishment of ICU mortality risk prediction models with machine learning algorithm using MIMIC-IV database^39^ | Journal | IV | TRUE | FALSE |
| Development and validation of a novel blending machine learning model for hospital mortality prediction in ICU patients with Sepsis^40^ | Journal | III | TRUE | FALSE |
| Machine learning algorithms for prediction of ventilator associated pneumonia in traumatic brain injury patients from the MIMIC-III database^41^ | Journal | III | TRUE | FALSE |
| Construction and validation of machine learning models for sepsis prediction in patients with acute pancreatitis^42^ | Journal | III & IV | TRUE | FALSE |
| A machine learning–based algorithm for the prediction of intensive care unit delirium (PRIDE): retrospective study^43^ | Journal | III | TRUE | TRUE |
| Dendrogram of transparent feature importance machine learning statistics to classify associations for heart failure: A reanalysis of a retrospective cohort study of the Medical Information Mart for Intensive Care III (MIMIC-III) database^44^ | Journal | III | TRUE | TRUE |
| Mortality prediction among ICU inpatients based on MIMIC-III database results from the conditional medical generative adversarial network^45^ | Journal | III | TRUE | TRUE |
| Machine learning prediction models for postoperative stroke in elderly patients: analyses of the MIMIC database^46^ | Journal | III & IV | FALSE | TRUE |
| Early predicting 30-day mortality in sepsis in MIMIC-III by an artificial neural networks model^47^ | Journal | III | TRUE | FALSE |
| Predicting mortality using machine learning algorithms in patients who require renal replacement therapy in the critical care unit^48^ | Journal | III | TRUE | FALSE |
| A simple weaning model based on interpretable machine learning algorithm for patients with sepsis: a research of MIMIC-IV and eICU databases^49^ | Journal | IV | TRUE | FALSE |
| Real-time mortality prediction using MIMIC-IV ICU data via boosted nonparametric hazards^50^ | Journal | IV | TRUE | FALSE |
| Critical correlation of predictors for an efficient risk prediction framework of ICU patient using correlation and transformation of MIMIC-III dataset^51^ | Journal | III | FALSE | TRUE |
| Machine Learning Approach to Predict Positive Screening of Methicillin-Resistant Staphylococcus aureus During Mechanical Ventilation Using Synthetic Dataset From MIMIC-IV Database^52^ | Journal | IV | TRUE | FALSE |
| Machine learning-based prediction of in-hospital mortality for critically ill patients with sepsis-associated acute kidney injury^53^ | Journal | IV | TRUE | TRUE |
| Explainable machine learning model for predicting furosemide responsiveness in patients with oliguric acute kidney injury^54^ | Journal | IV | TRUE | FALSE |
| Interpretable machine learning model for early prediction of 28-day mortality in ICU patients with sepsis-induced coagulopathy: development and validation^55^ | Journal | IV | TRUE | TRUE |
| Machine learning-based models for predicting mortality and acute kidney injury in critical pulmonary embolism^56^ | Journal | IV | TRUE | TRUE |
| Predicting risk for trauma patients using static and dynamic information from the MIMIC III database^57^ | Journal | III | TRUE | FALSE |
| Machine learning-based mortality prediction model for critically ill cancer patients admitted to the intensive care unit (CanICU)^58^ | Journal | III | TRUE | FALSE |
| Prediction of acute kidney injury in patients with liver cirrhosis using machine learning models: evidence from the MIMIC-III and MIMIC-IV^59^ | Journal | III & IV | TRUE | FALSE |
| Machine learning-based prediction model of acute kidney injury in patients with acute respiratory distress syndrome^60^ | Journal | III & IV | TRUE | TRUE |
| Early prediction of MODS interventions in the intensive care unit using machine learning^61^ | Journal | III & IV | TRUE | FALSE |
| Prediction of in-hospital mortality of intensive care unit patients with acute pancreatitis based on an explainable machine learning algorithm^62^ | Journal | IV | TRUE | FALSE |
| Prediction of in-hospital mortality for icu patients with heart failure^63^ | Journal | III | TRUE | TRUE |
| Practical machine learning-based sepsis prediction^64^ | Journal | III | TRUE | FALSE |
| 30-day hospital readmission prediction using MIMIC data^65^ | Journal | III | FALSE | TRUE |
| A Retrospective cohort study: predicting 90-day mortality for ICU trauma patients with a machine learning algorithm using XGBoost using MIMIC-III database^66^ | Journal | III | FALSE | TRUE |
| A predictive model for the risk of sepsis within 30 days of admission in patients with traumatic brain injury in the intensive care unit: a retrospective analysis^67^ | Journal | III & IV | TRUE | FALSE |
| Prostate cancer prediction model: a retrospective analysis based on machine learning using the MIMIC-IV database^68^ | Journal | IV | TRUE | TRUE |
| Predictive model of acute kidney injury in critically ill patients with acute pancreatitis: a machine learning approach using the MIMIC-IV database^69^ | Journal | IV | TRUE | TRUE |
| A Machine Learning pipeline using KNIME to predict hospital admission in the MIMIC-IV Database^70^ | Journal | IV | TRUE | FALSE |
| Comparison of machine learning algorithms for mortality prediction in intensive care patients on multi-center critical care databases^71^ | Journal | III & IV | TRUE | FALSE |
| Predicting in-hospital mortality for MIMIC-III patients: A nomogram combined with SOFA score^72^ | Journal | III | TRUE | TRUE |
| Machine Learning Model for the Prediction of Hemorrhage in Intensive Care Units^73^ | Journal | III & IV | TRUE | FALSE |
| Prediction of 30-day mortality for ICU patients with Sepsis-3^74^ | Journal | III | TRUE | FALSE |
| Development and validation of a machine-learning model for prediction of hypoxemia after extubation in intensive care units^75^ | Journal | IV | TRUE | TRUE |
| Machine learning as a tool to identify inpatients who are not at risk of adverse drug events in a large dataset of a tertiary care hospital in the USA^76^ | Journal | IV | TRUE | FALSE |
| Development and validation of a machine-learning model for predicting the risk of death in sepsis patients with acute kidney injury^77^ | Journal | IV | TRUE | TRUE |
| Survival Prediction in Patients with Hypertensive Chronic Kidney Disease in Intensive Care Unit: A Retrospective Analysis Based on the MIMIC^78^ Database | Journal | III | FALSE | TRUE |
| On the early detection of Sepsis in MIMIC-III^79^ | Journal | III | TRUE | FALSE |
| Comparison of machine learning algorithms to SAPS II in predicting in-hospital mortality of fractures of the pelvis and acetabulum: analyzes based on MIMIC-III database^80^ | Journal | III | TRUE | FALSE |
| Predicting Mortality in Sepsis-Associated Acute Respiratory Distress Syndrome: A Machine Learning Approach using the MIMIC-III database^81^ | Journal | III | TRUE | FALSE |
| Explainable Machine Learning Models for Pneumonia Mortality Risk Prediction Using MIMIC-III Data^82^ | Journal | III | TRUE | TRUE |
| Simplified &Novel Predictive Model using Feature Engineering over MIMIC-III Dataset^83^ | Journal | III | TRUE | TRUE |
| XGBoost in the Prediction of 28-Day Mortality in Critical Elderly Patients with Hip Fracture: A MIMIC-IV Cohort Study^84^ | Preprint | IV | TRUE | TRUE |
| Predicting Prescribed Medications from the MIMIC-IV Event and Measurement Data^85^ | Journal | IV | TRUE | FALSE |
| Clinical nomogram prediction model to assess the risk of prolonged ICU length of stay in patients with diabetic ketoacidosis: a retrospective analysis based on the MIMIC-IV database^86^ | Journal | IV | TRUE | FALSE |
| Internal and external validation of machine learning–assisted prediction models for mechanical ventilation–associated severe acute kidney injury^87^ | Journal | IV | TRUE | TRUE |
| Nomogram establishment for short-term survival prediction in ICU patients with aplastic anemia based on the MIMIC-IV database^88^ | Journal | IV | TRUE | TRUE |
| Predicting patient outcome from clinical journals and biomedical articles: Using the MIMIC-IV database, multiple in-hospital mortality prediction models are created, to which improvements are attempted through the use of word embeddings trained on scientific biomedical literature^89^ | Journal | IV | TRUE | FALSE |
| Machine learning time-to-event mortality prediction in MIMIC-IV critical care database^90^ | Journal | IV | TRUE | FALSE |
| Machine learning models to predict 30-day mortality for critical patients with myocardial infarction: a retrospective analysis from MIMIC-IV database^91^ | Journal | IV | TRUE | NA |
| ML-Based AKI Prediction in Acute Pancreatitis: Innovative Models from MIMIC-IV Database^92^ | Preprint | IV | TRUE | TRUE |
| Exploring Predictive Factors for Heart Failure Progression in Hypertensive Patients Based on Medical Diagnosis Data from the MIMIC-IV Database^93^ | Journal | IV | FALSE | TRUE |
| A novel clinical prediction model for in-hospital mortality in sepsis patients complicated by ARDS: A MIMIC IV database and external validation study^94^ | Journal | IV | TRUE | TRUE |
| Factors and machine learning models for predicting successful discontinuation of continuous renal replacement therapy in critically ill patients with acute kidney injury: a retrospective cohort study based on MIMIC-IV database^95^ | Journal | IV | TRUE | NA |
| Machine learning for in-hospital mortality prediction in critically ill patients with acute heart failure: A retrospective analysis based on MIMIC – IV databases^96^ | Preprint | IV | TRUE | TRUE |
| Evaluating the Fairness of the MIMIC-IV Dataset and a Baseline Algorithm: Application to the ICU Length of Stay Prediction^97^ | Preprint | IV | TRUE | NA |
| Multitask learning to predict successful weaning in critically ill ventilated patients: A retrospective analysis of the MIMIC-IV database^98^ | Journal | IV | TRUE | NA |
| Development of machine learning models for predicting acute respiratory distress syndrome: evidence from the MIMIC-III and MIMIC-IV^99^ | Preprint | III & IV | TRUE | NA |
| Feature selection and risk prediction for diabetic patients with ketoacidosis based on MIMIC-IV^100^ | Journal | IV | TRUE | TRUE |
